# Using multiomic data to predict postoperative complications after major surgery in the UK Biobank cohort

**DOI:** 10.64898/2026.03.10.26348039

**Authors:** Richard A. Armstrong, Paul Yousefi, Ben Gibbison, Golam M Khandaker, Tom R Gaunt

**Affiliations:** MRC Integrative Epidemiology Unit, Bristol Medical School, University of Bristol, Bristol, UK; NIHR Bristol Biomedical Research Centre, University Hospitals Bristol and Weston NHS Foundation Trust and University of Bristol, Bristol, UK; Bristol Medical School, University of Bristol, Bristol, UK; Avon and Wiltshire Mental Health Partnership NHS Trust, Bristol, UK; Centre for Academic Mental Health, Population Health Sciences, Bristol Medical School, University of Bristol, Bristol, UK

**Keywords:** biobank, multiomics, perioperative risk prediction, postoperative complications

## Abstract

**Background:** Whilst high-dimensional omic profiles from biobanks can predict long-term disease outcomes, their ability to predict postoperative outcomes is unknown. We investigated whether adding metabolomic and proteomic data from historic samples to standard clinical variables improved the prediction of postoperative complications.

**Methods:** We analysed data from 158,156 UK Biobank participants who underwent major surgery. The primary outcomes were postoperative atrial fibrillation, acute kidney injury, acute myocardial infarction, delirium, stroke and surgical site infection. We developed machine learning models (elastic net penalised regression) to compare baseline clinical variables against integrated single- and multiomic datasets. To address sample size constraints in high-dimensional omic subsets, we employed transfer learning from related non-postoperative domains.

**Results:** The numbers of cases with omic data varied across outcome phenotypes and feature sets: metabolomic: 144–1596, proteomic: 27–289 and multiomic: 15–219. Baseline clinical models achieved robust predictive performance (AUROC 0.72– 0.88, sensitivity 0.71–0.80). The addition of metabolomic and/or proteomic features provided no clinically meaningful improvement in performance for any clinical phenotypes. Transfer learning from the non-postoperative domain improved model performance and stability but did not outperform baseline clinical models.

**Conclusions:** The addition of metabolomic and proteomic data from samples collected at a temporal distance from surgery does not improve preoperative risk prediction compared to standard clinical variables. The lack of incremental predictive value suggests that long-term biological trajectories have become manifest in routine clinical data by the time of surgery. The success of transfer learning from non-postoperative settings may indicate shared biological risk between chronic and acute phenotypes.

## Introduction

Postoperative complications are a major driver of morbidity, mortality and resource utilisation, occurring in up to 15% of patients worldwide ^1^, and can have serious long-term implications for patients ^2–4^. Current preoperative risk stratification tools rely heavily on demographic factors and clinical scoring systems, for example ASA physical status grade ^5, 6^. However, these fail to capture the biological heterogeneity of individual patients and typically leave a substantial component of perioperative risk unaccounted for ^7^.

One approach to add granularity is the use of high-dimensional molecular assays which provide snapshots of the interplay between genetic predisposition, environmental or lifestyle factors, and subclinical disease states ^8, 9^. In other contexts, metabolomic and proteomic signatures from large-scale biobanks have been shown to act as markers of underlying health or multimorbidity trajectories, preceding clinical diagnoses and predicting long-term outcomes ^10, 11^. They might therefore provide incremental predictive value over standard clinical variables for postoperative outcomes, even when sampled months to years before surgery occurs.

However, as the number of measured biomarkers increases, the curse of dimensionality becomes an additional limitation, particularly with limited sample sizes. One approach to counter this is dimensionality reduction, for example through the use of principal component analysis (PCA) or autoencoders (AEs) ^12^, which can be used to limit the number of features to a lower dimension space. An alternative approach is transfer learning, in which prediction patterns are learned from a related source domain with more abundant data and the resulting models are applied or fine-tuned in the target domain ^13^.

To explore the use of multiomics in preoperative risk stratification we therefore defined six clinically important postoperative phenotypes, atrial fibrillation (AF), acute kidney injury (AKI), acute myocardial infarction (AMI), delirium, surgical site infection (SSI) and stroke, in a large cohort of patients undergoing major surgery in the UK Biobank. We investigated whether the addition of metabolomic and/or proteomic data to baseline clinical predictors improved predictive accuracy and whether transfer learning from related, non-postoperative phenotypes could overcome the limitations of previous studies.

## Methods

### Study population

UK Biobank is a large-scale biomedical database and research resource containing genetic, lifestyle and health information from half a million UK participants recruited between 2006 and 2010 (http://www.ukbiobank.ac.uk). Linked health data is available for all participants with follow-up to 2022 ^14^. The UK Biobank study was approved by the North-West Multi-centre Research Ethics Committee (reference 21/NW/0157) and all participants provided written informed consent. This research has been conducted under project number 128619.

### Metabolomic and proteomic assays

A subset of UK Biobank participants had metabolomic and/or proteomic assay data available at the time of analysis. Metabolic biomarkers were measured from randomly selected EDTA plasma samples using a high-throughput NMR-based metabolic biomarker profiling platform developed by Nightingale Health Ltd. Proteomic analysis was performed using the Olink Explore platform, a Proximity Extension Assay (PEA). Further details on sample selection and quality control procedures are available from UK Biobank ^15, 16^.

### Overview of modelling approaches

The primary analysis used data from the UK Biobank surgical cohort (defined below) to predict the postoperative complications of interest (Figure 1, ‘direct postoperative modelling’). First, clinical covariates were used to train baseline clinical models for each outcome. Combined clinical and omic models were then trained for each outcome and omic feature set (metabolomics, proteomic and multiomics). The transfer learning analysis used data from the non-surgical UK Biobank cohort to predict non-postoperative outcomes (Figure 1, ‘transfer learning’). Models were trained using clinical covariates with the addition of metabolomic, proteomic and multiomic data for each non-postoperative outcome. Then, to ‘transfer’ the learning, the resulting models were applied to the surgical cohort to generate a risk prediction for each non-postoperative outcome. This predicted risk was then used as a feature in the final transfer models to predict the relevant postoperative complications. For both direct and transfer learning, omic-only models were also trained for each outcome and omic feature set (without clinical covariates) to assess the inherent biological signal in the omic datasets and validate the transfer learning approach.

**Figure 1.**
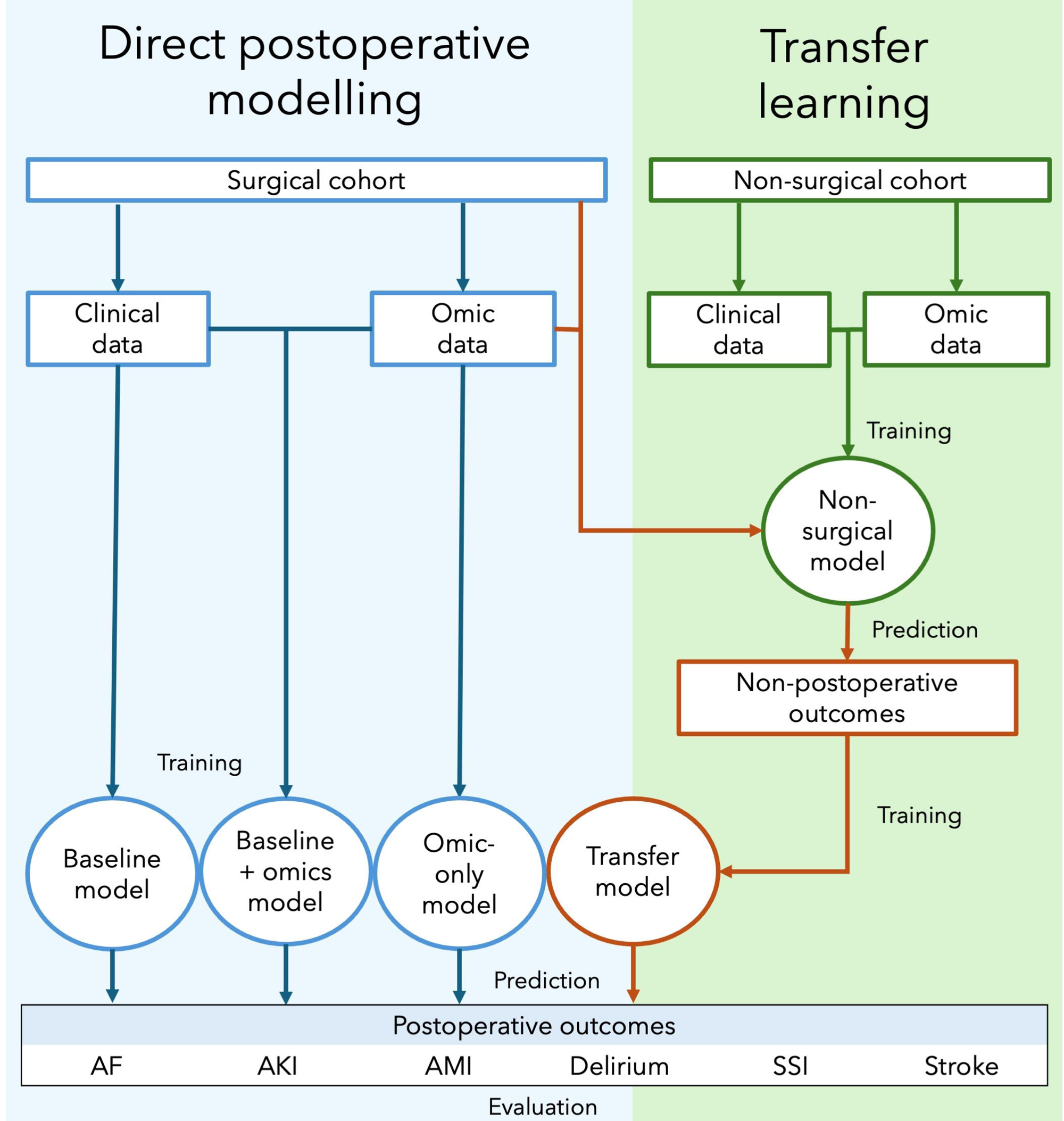
Schematic representation of study pipeline. Direct postoperative modelling (blue) represents using the surgical UK Biobank cohort to train predictive models based on the postoperative outcomes of interest. Transfer learning (green) represents using the non-surgical UK Biobank cohort to train predictive models based on the non-postoperative equivalent outcome phenotypes. The surgical cohort are then passed through the non-surgical model and a predicted risk for the non-surgical outcome is used as a feature in a transfer model to predict the postoperative outcome (orange).

### Direct postoperative modelling: inclusion and exclusion criteria

For direct postoperative modelling, eligible participants were those with metabolomic and/or proteomic data available who also underwent major, inpatient surgery after their date of enrolment in UK Biobank. Major surgery was defined by OPCS4 codes using the Bupa Schedule of Procedures and Abbott classification ^17–19^. For AF, AKI, AMI and SSI, participants aged 18 and over at the time of surgery were included. For postoperative delirium and stroke, we restricted the analysis to those aged 60 or over. Exclusion criteria for all phenotypes included a previous diagnosis of the outcome of interest and planned day case surgery (see Supplementary Methods 1 for more detail on phenotype definition and exclusions).

### Direct postoperative modelling: outcome definition

The primary outcome for each complication was a first diagnosis of that condition within 30 days of surgery. Controls were defined as those meeting the inclusion criteria but without the complication occurring within 30 days of surgery. Cases and controls were defined separately for each outcome phenotype using the following International Classification of Diseases, Tenth Revision (ICD-10) codes: atrial fibrillation, I48.9; acute kidney injury, N17; acute myocardial infarction, I21; delirium, F05; stroke, I61/I63/I64; and surgical site infection, T81.4.

### Direct postoperative modelling: modelling dataset

The modelling dataset included clinical covariates, metabolomic assay data and proteomic assay data, with the outcome phenotype considered as a binary variable. Clinical covariates were selected to reflect those available to clinicians in routine clinical practice: age, sex, Charlson Comorbidity Index (CCI, coded as a binary variable 0-1 vs ≥2 ^20^), admission method (elective or emergency) and operation category (major or complex). Metabolomic data included the 251 measured biomarkers in the Nightingale panel ^16^ plus the spectrometer (1-9) used for analysis. The Olink proteomic data is comprised of multiple panels ^15^. Models were developed using the Inflammation I panel (368 biomarkers), the combination of Inflammation I and Inflammation II (737 biomarkers), and all proteins (2922 biomarkers). Table 1 shows the feature sets used for modelling.

**Table 1.**
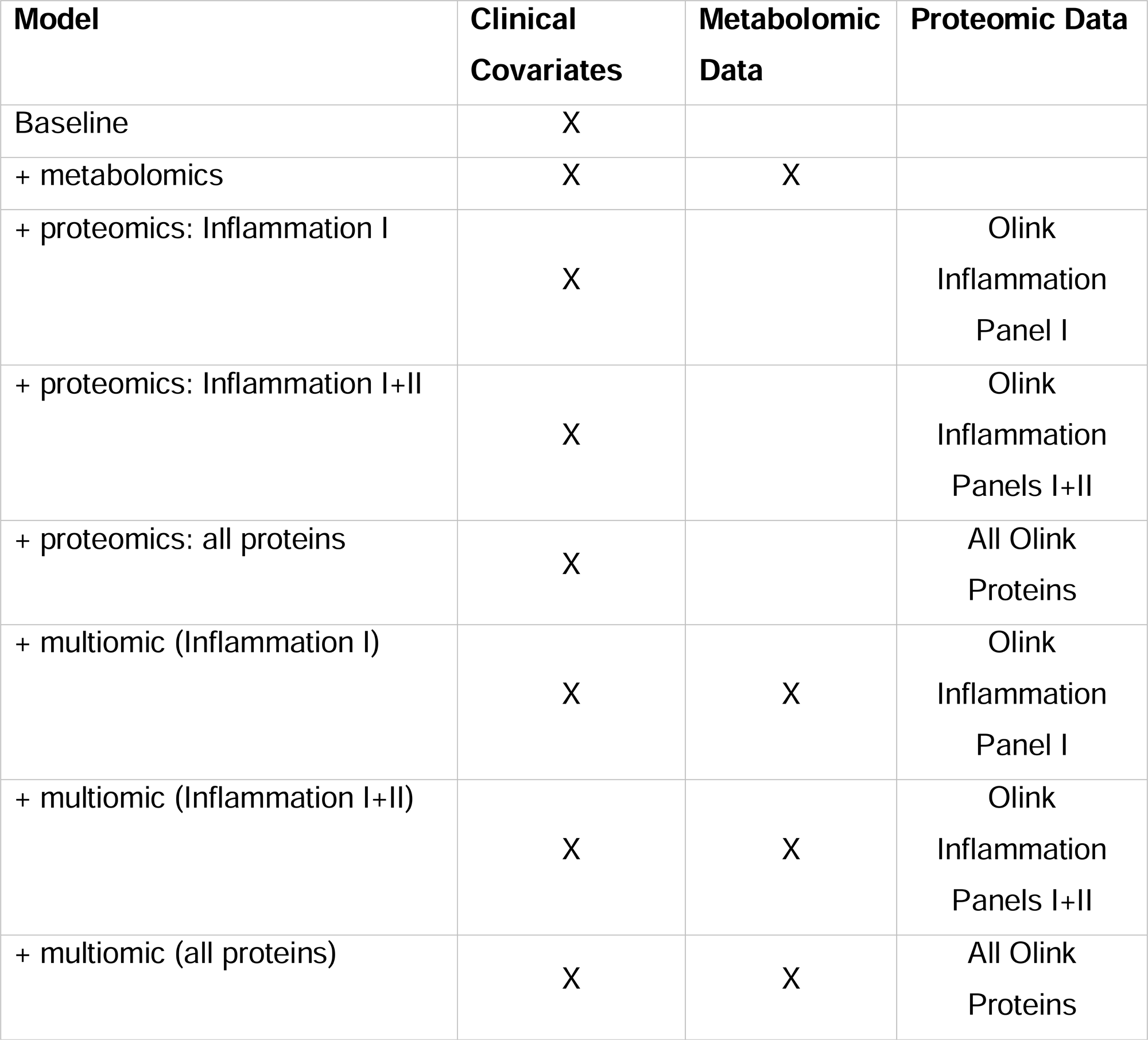
Feature sets used in direct postoperative modelling.

### Transfer learning: inclusion and exclusion criteria

For transfer learning, the non-surgical subset of UK Biobank (i.e. those participants with metabolomic and/or proteomic data available who did not undergo major, inpatient surgery after their date of enrolment in UK Biobank) were used as the source domain to prevent any sample overlap with the postoperative cohort.

### Transfer learning: outcome definition

For each postoperative complication, a chronic or non-postoperative equivalent phenotype was defined, for example chronic atrial fibrillation or non-postoperative stroke, using the same ICD-10 codes. For surgical site infection, an alternative diagnosis of ‘infections of the skin and subcutaneous tissue’ (ICD-10 codes L00-L08) was used.

### Transfer learning: modelling dataset

The modelling dataset included clinical covariates, metabolomic assay data and proteomic assay data, with the outcome phenotype considered as a binary variable. Clinical covariates included age, sex and CCI, to align with the postoperative analyses. For cases, age and CCI were calculated at the time of the hospital admission related to the diagnosis. To avoid immortal time bias, incidence density sampling was used to assign follow-up times to controls ^21^. In brief, the time interval from UK Biobank enrolment to diagnosis for each case was used to generate a distribution of potential follow-up durations. For each control participant, a follow-up duration was randomly assigned from this distribution with age and CCI taken from the most recent hospital episode at that point (i.e. date of UK Biobank enrolment plus follow-up duration). A case-control ratio of 1:4 was used and models were developed using the same feature sets as in the direct postoperative modelling (Table 1).

### Data preprocessing and dimensionality reduction

Data were preprocessed prior to model development. Variables that were missing in ≥10% of participants were omitted, and individuals with missing data for ≥80% of features were excluded. Clinical numerical variables were centred and scaled after imputation of missing values using the median. Categorical data were one-hot encoded with missing values imputed with a constant (‘missing’).

Multiomic data were prepared for analysis using the metaboprep pipeline (v1.0 ^22^) with default parameters. Metabolomic data were logit (for percentages) or log (for other biomarkers) transformed then centred and scaled. Missing values were imputed with the biomarker mean (0). For proteomic data, which is provided by Olink as log_2_ normalised protein expression (NPX) values, missing values were imputed with the biomarker mean then rank-based inverse normal transformation was applied. Finally, both metabolomic and proteomic features were clipped at ±10 standard deviations before being passed through a custom autoencoder (AE) for dimension reduction. The AEs were designed as symmetrical ‘bottleneck’ networks, functioning as non-linear feature extractors to compress omic inputs into a 16-dimensional latent space. Architectures were determined through an iterative empirical search, evaluating performance based on reconstruction loss (minimising mean squared error between input and output, using Principal Component Analysis (PCA) as a baseline benchmark) and generalisability (monitoring the divergence between training and validation loss to limit overfitting). To accommodate the varying dimensionality of different omic feature sets, the depth and width of hidden layers were scaled based on the input data (full specifications for each feature set are in Supplementary Methods 2). The AEs were trained for 20 epochs using the Adam optimiser with a learning rate of 10^-3^ and batch size of 32. For multiomic models (those including both metabolomic and proteomic features) both early and late integration approaches were explored. Early integration combined metabolomic and proteomic data into a single AE; late integration trained separate AEs for metabolomic and proteomic data with the outputs of both entering the classifier models (Supplementary Figure 1).

### Model training: baseline and omic models

For direct postoperative modelling (baseline, combined and omic-only models) the Elastic Net logistic regression algorithm was used for model development as it facilitates simultaneous variable selection and model regularisation by combining LASSO (L1) and ridge (L2) penalty terms. To address the significant class imbalance, we trained the Elastic Net models within a Balanced Bagging Classifier. Each bagging classifier trained 50 base estimators (Elastic Net models) on different randomly selected subsets of the available data, undersampling the majority class to achieve a 1:1 case-control ratio (‘balanced’). The predicted probabilities of each base estimator were then averaged to provide the final prediction (‘bagging’). This ensemble approach stabilises the coefficient estimates and ensures the model is not biased toward the majority class.

To obtain unbiased performance estimates, the bagging classifiers were trained within an adaptive repeated stratified k-fold cross-validation framework (Supplementary Figure 2). *k*-fold cross-validation splits the dataset into *k* partitions (folds). For each fold, models are trained using the other *k-1* folds and tested on the held-out fold. In repeated *k*-fold cross-validation, this process is repeated multiple times with different splits of the data on each repeat. The average performance across all folds then provides a less biased estimate of generalisation performance. The number of folds and repetitions was dynamically adjusted based on the number of positive cases (*N*) for each complication and feature set (20 repeats of 3-fold cross-validation for *N* <100; 12 repeats of 5-fold cross-validation for *N* ≥100). Where *N* ≥500, an additional 3-fold inner cross-validation loop was used for hyperparameter tuning (*C* and *L1 ratio*; full grid in Supplementary Methods 3). For *N* <500, no inner cross-validation was performed and hyperparameter values were fixed (*C* 1.0; *L1 ratio* 0.5).

To ensure equitable comparison, a distinct clinical baseline model was developed for each outcome-omic dataset combination (due to differences in available sample size) using identical training/testing partitions, maintained via a fixed random seed. Baseline models included age, sex, CCI, operative category and admission method. Combined omic models included these clinical variables, plus the relevant omic feature set.

### Model training: transfer model

To transfer from the source (non-postoperative) to the target (postoperative) domain final trained models for each non-postoperative phenotype-dataset combination were used to generate a predicted probability of that outcome for each postoperative participant. The inputs to each model were age, sex, CCI and the relevant omic data. The resulting predicted probabilities were then used alongside admission type and operative category (as they are not applicable in the non-postoperative domain and so not included in the non-postoperative models) to train parsimonious ‘transfer’ models predicting the postoperative outcomes of interest (Figure 1 and Supplementary Methods 4). These reduced models were trained using logistic regression with an L2 penalty term in the same balanced bagging classifier cross-validation framework described above.

### Model evaluation

Performance metrics were calculated for each of the 60 outer cross-validation folds (either 20 x 3-fold cross-validation or 12 x 5-fold cross-validation) for each model to estimate generalisation performance. To reflect the potential clinical usage of these models in identifying those at increased risk of complications the primary metrics reported are area under the receiver operating characteristic curve (AUROC), recall (sensitivity) and specificity (calculated as 2*balanced accuracy – recall) at the standard probability threshold of 0.5. To assess the incremental benefit of the added omics features we compared the performance of each combined omics model against its related baseline model using the Corrected Resampled t-test ^23^. This approach was selected to account for the lack of independence between folds in our repeated cross-validation design, which otherwise leads to an underestimation of variance and an inflated Type I error rate. To account for multiple hypothesis testing across the different feature sets and model comparisons, p-values were adjusted using the Benjamini-Hochberg False Discovery Rate (FDR) procedure, with statistical significance defined as a FDR < 0.05. Corrections were applied separately for each performance metric (AUROC, recall, specificity) and each clinical outcome (AF, AKI, delirium, AMI, stroke, SSI), as these represent distinct groups of clinical hypotheses. A post-hoc assessment of sample size was made for each outcome-feature set combination with reference to guidelines on required sample size for predictive modelling (Supplementary Methods 5) ^24^.

Analyses were performed in Python (3.6.8) with scikit-learn (0.24.2) and imbalanced-learn (0.8.1) for preprocessing, Elastic Net and Balanced Bagging Classifier models, tensorflow/keras (2.6.2) for autoencoder classes, and scipy.stats (1.5.4) and statsmodels (0.12.2) for model evaluation.

## Results

### Direct postoperative modelling: participant characteristics

The UK Biobank dataset contained 158,156 individuals undergoing qualifying surgery of whom 4743 had omic data available and experienced at least one of the postoperative complications of interest (Table 2). Post-hoc power differed across postoperative complications as the proportion of individuals with metabolomic, proteomic and both datasets varied (Supplementary Tables 1-2). For metabolomic data, case numbers ranged from 144 for AMI to 1596 for AKI; proteomics were available for between 27 and 289 cases; and multiomic data were present for between 15 and 219 individuals. The median (IQR [range]) time between sampling and surgery was 6 (2.9–9.4 [1 day – 15.5]) years.

**Table 2.**
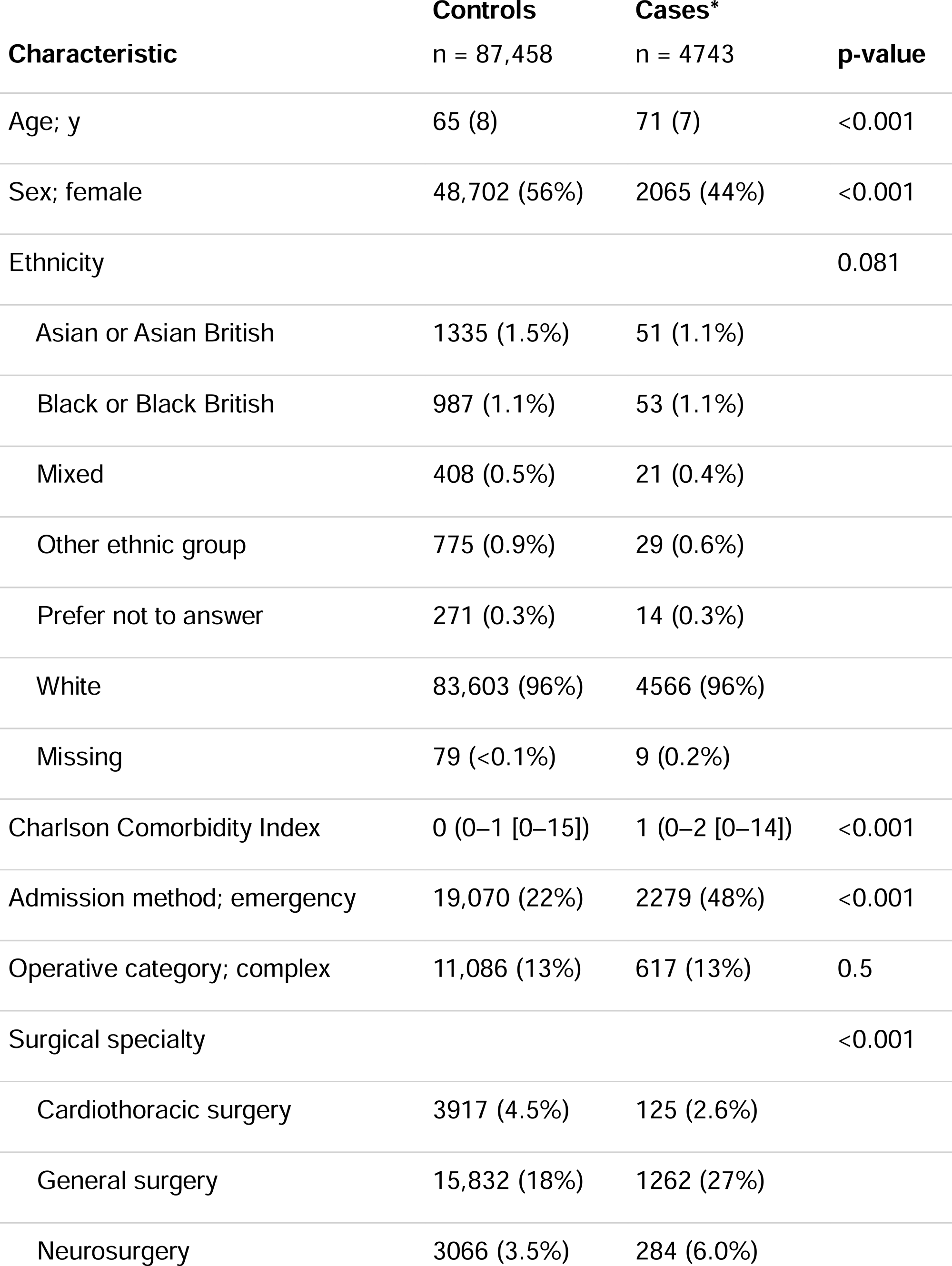
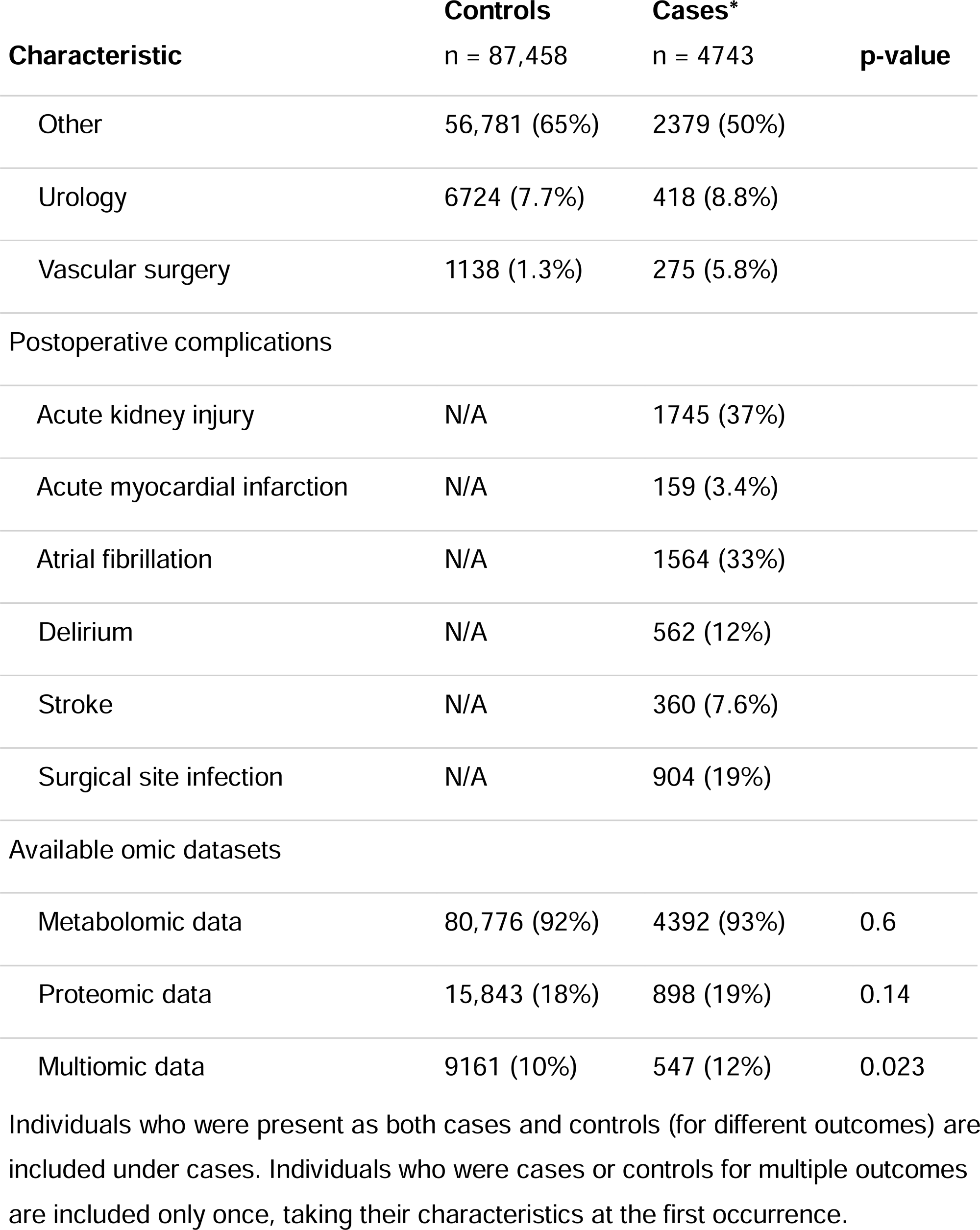
Participant characteristics of individuals undergoing major surgery in UK Biobank with available metabolomic and/or proteomic data. Values are mean (SD), median (IQR [range]) or number (proportion).

### Direct postoperative modelling: baseline models

Distinct baseline models were trained for the subset of participants who had available data for each of the omic feature sets. Overall performance of the baseline models was good, though feature sets with smaller sample sizes (multiomics) tended to show greater variability (Figure 2, Panels A–F).

**Figure 2.**
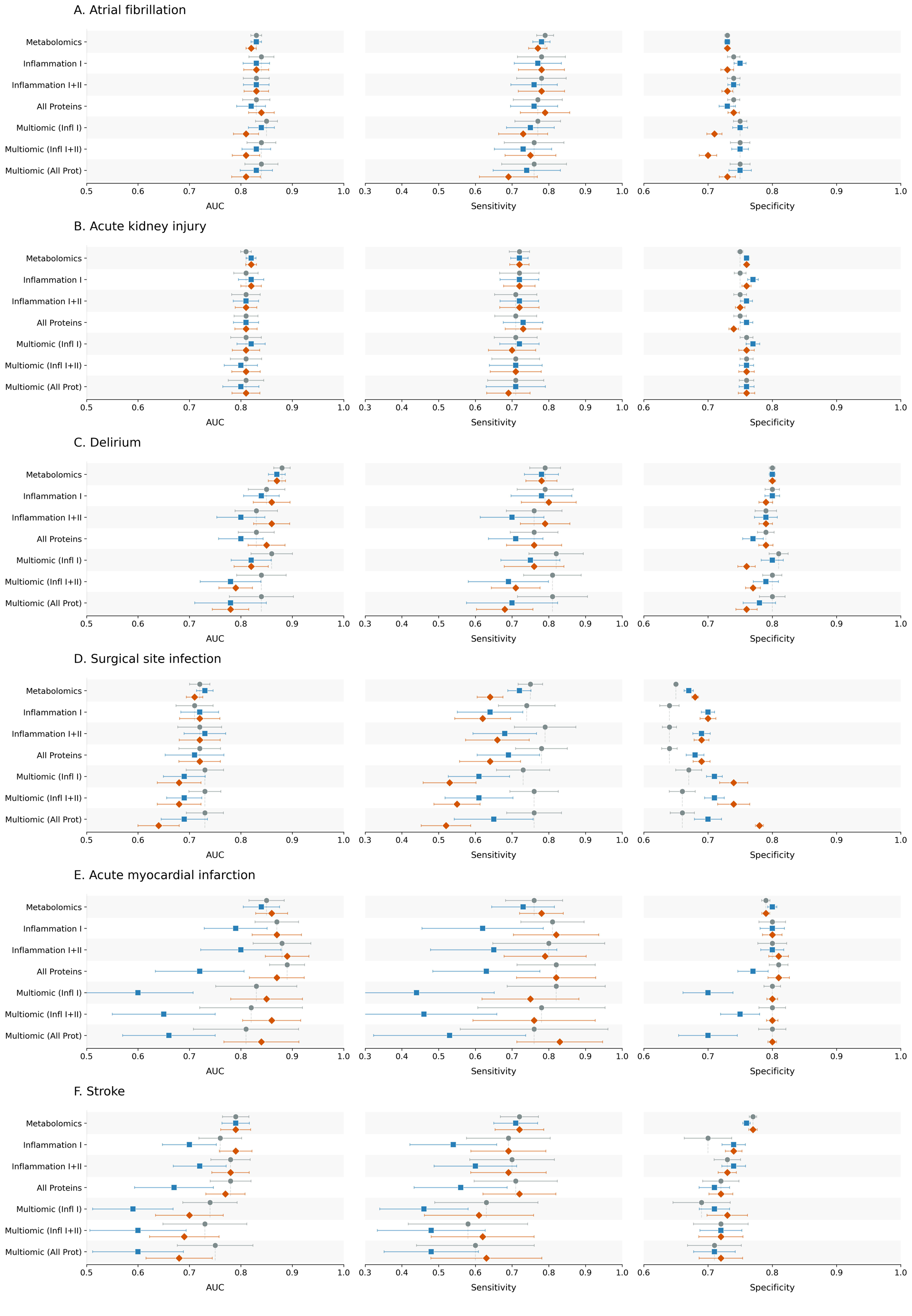
Evaluation metrics for each postoperative complication and feature set. Baseline models (grey circles 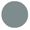); omics models (blue squares 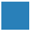); transfer models (red diamonds 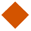). Values are mean with standard deviation error bars. AUC: area under the receiver operator characteristic curve.

### Direct postoperative modelling: omic-only models

Omic-only models demonstrated modest but consistent predictive ability across most outcome phenotypes, confirming the presence of a biological signal in the omic data despite temporal distance from the surgical event (Supplementary Table 3). The strongest standalone signals were observed for postoperative AKI using metabolomic and/or inflammation I proteins (AUC (SD) metabolomics 0.63 (0.01); inflammation I 0.67 (0.03); metabolomics and inflammation I 0.66 (0.03)). Stroke and AMI tended to perform less well but the sample sizes for these outcomes were below recommended levels for model development (Supplementary Table 2)

### Direct postoperative modelling: metabolomics

Baseline models trained in the subset of participants with metabolomic data achieved good performance with AUROC ranging from 0.72 (± 0.02) to 0.88 (± 0.02) and sensitivity from 0.71 (± 0.06) to 0.78 (± 0.02). Across all outcomes, adding metabolomic features to the baseline clinical models did not produce clinically-meaningful improvements in any performance metrics. Delirium and AF showed the best performance with models for SSI tending to show the worst performance (Figure 2, Panels A–F; Supplementary Tables 4.1–4.6). Results for the AMI metabolomic model showed greater variability which is reflective of the sample size being below recommended levels (Supplementary Table 2).

### Direct postoperative modelling: proteomics

In the proteomic cohort, baseline clinical models performed well for AF, AKI and delirium (AUROC 0.81–0.85; sensitivity 0.71–0.80) and performance was not improved by the addition of proteomic features (Figure 2, Panels A–C; Supplementary Tables 4.1–3.3). Models for SSI tended to perform least well (AUC 0.71–0.72, sensitivity 0.74–0.79) though specificity was improved by the addition of proteomic features (inflammation I 0.64 (± 0.02) vs 0.7 (± 0.01), inflammation I+II 0.64 (± 0.01) vs 0.69 (± 0.01), all proteins 0.64 (± 0.01) vs 0.68 (± 0.01), all p < 0.001 for corrected resampled t-test comparing 60 cross-validation folds; Figure 2, Panel D, Supplementary Table 4.5). Sample sizes for stroke and AMI were less than 33% of recommended levels so were not considered further for proteomic modelling (Supplementary Table 2).

### Direct postoperative modelling: multiomics

In the multiomic cohort, baseline models for AF and AKI showed good performance (AUC 0.81–0.85, sensitivity 0.71–0.77, specificity 0.75–0.76) and were not improved by the addition of multiomic features by early integration (Figure 2, Panel A–B; Supplementary Tables 4.1–4.2). Sample sizes for the other outcomes and for all outcomes using the late integration approach were substantially lower than recommended levels so not considered further (Supplementary Table 2).

### Transfer learning

Transfer learning with paired phenotypes was used to address the sample size constraints and resulted in case numbers of between 3.4 and 12.5 times those of the target (postoperative) cohort, depending on phenotype and feature set (Supplementary Table 5). This resulted in improved performance and stability in the omic-only models, particularly for outcomes with limited postoperative sample sizes, confirming a valid biological signal within the historic omic data (Supplementary Table 3).

The metabolomic dataset had the largest sample size for every outcome studied, meeting sample size recommendations for all except AMI (Supplementary Table 2). For AMI, transfer learning tended to improve model performance and reduce the variability of predictions, though no metrics showed statistically significant improvements across the cross-validation procedure (Figure 2, Panel E; Supplementary Table 4.4). For SSI, where an alternative diagnosis was used, transfer learning resulted in reduced sensitivity compared to baseline and direct learning models (transfer model 0.64 ± 0.04, baseline 0.75 ± 0.03, p < 0.001; direct 0.72 ± 0.03, p = 0.01, across 60 cross-validation folds) (Figure 2, Panel D, Supplementary Table 4.5).

For proteomic feature sets, sample sizes were below recommended levels for all outcomes except AKI and AF (Supplementary Table 2). Transfer learning tended to result in improved stability compared to models trained in the postoperative cohort but performance was similar (Figure 2, Panels C–F; Supplementary Tables 4.3–4.6).

For transfer models using multiomic feature sets (early integration), model stability was generally improved by transfer learning compared to models learned directly from the postoperative dataset, particularly for AMI and stroke, but did not result in statistically significant performance gains over clinical baseline models (Figure 2, Panels A–F; Supplementary Tables 4.1–4.6).

## Discussion

In this large-scale analysis of the UK Biobank cohort, we found that the addition of high-dimensional metabolomic and proteomic data to standard clinical variables produced no incremental benefit in predicting a broad range of postoperative complications. These findings remained consistent across six distinct clinical phenotypes, single- and multiomic datasets and multiple statistical approaches, despite the presence of biological signal within the historic omic samples. We also found that transfer learning from non-postoperative to postoperative domains produced good model performance whilst countering the limitations of small sample sizes.

A challenge presented when biobank data are used for perioperative risk prediction is the temporal gap between sample collection and the surgical event, which was a median of six years in this cohort. Metabolomic and proteomic assays provide dynamic snapshots of physiological homeostasis, reflecting the interplay between intrinsic and environmental factors ^8^. Historic samples may therefore not accurately reflect the physiological reserve available at the time of a particular surgical episode ^25^. For these biomarkers to add predictive value in the perioperative setting, they may need to be measured closer to the time of surgery to capture the immediate physiological context.

We did, however, see consistent discrimination performance in the omic-only models, particularly those trained in the non-postoperative domain, suggesting that the molecular trajectories captured by these assays do persist over time. The lack of incremental gain in our combined models, coupled with the robust predictive power of the baseline clinical models, might therefore indicate that the biological signal is not absent from the omic data, but rather it has already become manifest in the chronic comorbidities or age-related physiological decline captured by the clinical covariates. Whilst historic omic profiles may capture latent, undiagnosed pathology that more accurately reflects an individual’s true underlying multimorbidity, any potential predictive advantage is likely attenuated by the interval between sampling and surgery. This contrasts with biobank studies in the non-surgical setting, where omic assays successfully predict long-term chronic disease risk ^10, 11^. In those instances, omics are used to forecast future onset of disease, whereas in this study we are essentially looking backwards from the point of surgery. If the biological risk has already culminated in a diagnosis or physiological decline, the high-resolution assay provides little additional information over up-to-date clinical records.

To address the constraints of modelling rare events with high-dimensional inputs and limited sample sizes, we applied transfer learning from the non-postoperative domain. This resulted in improved performance relative to the models trained in the original target domain, particularly in terms of model stability (as would be expected with a larger effective sample size). From a biological perspective, the success of this transfer learning suggests that the paired postoperative and non-postoperative phenotypes have a shared underlying architecture of clinical and omic risk factors.

The systemic inflammatory stress of major surgery may therefore accelerate the appearance of a phenotype in an individual with biological predisposition: rather than representing *de novo* pathologies in the postoperative period, these complications may represent an acute unmasking of latent vulnerabilities in at-risk individuals.

Whether these phenotypes would have become manifest without surgery as a ‘trigger’ cannot be determined from these analyses. Of note, we used an alternative diagnosis as the source domain for SSI and saw decreased sensitivity when predicting the postoperative outcome. This highlights the limitations of the approach and the importance of selecting an appropriate context from which to transfer learning.

From a translational perspective, these findings support the principle of parsimony in preoperative assessment and risk stratification. Existing scoring systems based on demographic and clinical inputs are easily accessible, resource-efficient and interpretable ^26, 27^. Our baseline models achieved robust discrimination despite using only five fundamental variables and lacking near-ubiquitous predictors such as ASA physical status grade. This suggests that a small number of clinical factors capture the majority of predictable risk, leaving little capacity for improvement from additional high-throughput omics collected at a temporal distance from surgery. However, it remains possible that omic data would provide significant incremental benefit if measured in the immediate perioperative period, capturing time-sensitive preclinical physiological decline that has not yet resulted in a formal diagnosis. Further research should explore this approach as a means of deriving benefit from the additional resolution these assays provide.

Strengths of this study include the use of a large, deeply-phenotyped cohort and a rigorous methodological pipeline to prevent data leakage, reduce the risk of overfitting and provide unbiased estimates of model performance. Additionally, by evaluating distinct outcome phenotypes across organ systems we demonstrated that the lack of positive findings was not outcome-specific. However, the study does have limitations. The UK Biobank cohort are not representative of the wider population due to a ‘healthy volunteer’ bias ^28^, which may affect generalisability. Postoperative outcomes were derived from clinical coding data and were not prospectively screened for, so underreporting is likely. Some clinical variables which are widely used in existing risk prediction tools are not present in the linked healthcare data (e.g. ASA physical status grade, type of anaesthesia, surgical urgency) and so could not be included. Finally, despite the large overall size of the UK Biobank cohort, the sample size for the specific outcomes studied and the fact that only a proportion of individuals had omic profiling limited our statistical power to detect small effect sizes.

Overall, this study demonstrates that whilst metabolomic and proteomic profiles measured at a temporal distance from surgery contain persistent biological signals, they do not provide incremental value over standard clinical variables for preoperative risk stratification. The additional resolution provided has likely become manifest over time and so the additional cost and complexity of historic omic profiling is not justified. However, given the success of the transfer learning approach employed, future work using larger cohorts of non-postoperative patients may produce insights which can be applied to the perioperative setting. Alternatively, multiomic sampling closer to the surgical event may add additional benefit as it can capture the immediate physiological context in which the inflammatory insult of major surgery occurs.

## Acknowledgements

This research has been conducted using the UK Biobank Resource under Application Number 128619. This includes linked data provided by patients and collected by the NHS as part of their care and support and data assets made available by National Safe Haven as part of the Data and Connectivity National Core Study, led by Health Data Research UK in partnership with the Office for National Statistics and funded by UK Research and Innovation (grant ref MC_PC_20058). This work was carried out using the computational and data storage facilities of the Advanced Computing Research Centre, University of Bristol (http://www.bristol.ac.uk/acrc/).

This study was funded by a Wellcome Trust GW4-CAT PhD Programme for Health Professionals PhD Fellowship awarded to RAA [316275/Z/24/Z]. RAA, PY, GMK and TRG are supported by the Medical Research Council Integrative Epidemiology Unit at the University of Bristol (RA, TG: MC_UU_00032/3; PY: MC_UU_00032/4; GMK: MC_UU_0032/6). GMK acknowledges additional funding from the Wellcome Trust (grant numbers: 201486/Z/16/Z and 201486/B/16/Z), the Medical Research Council (grant numbers: MR/W014416/1; MR/S037675/1; MR/Z50354X/1; and MR/Z503745/1. BG, TRG, GMK and PY are also supported by the UK National Institute for Health and Care Research (NIHR) Bristol Biomedical Research Centre (grant number: NIHR 203315). The views expressed are those of the authors and not necessarily those of the UK NIHR or the Department of Health and Social Care. The funders had no role in study design, data collection and analysis, decision to publish, or preparation of the manuscript.

## Declaration of interests

TRG receives funding from GlaxoSmithKline, Biogen, Novartis and Roche for unrelated research. No other competing interests declared.

## Data availability statement

The source data for these analyses are available from UK Biobank. Details of how to access UK Biobank data can be found here: https://www.ukbiobank.ac.uk/use-our-data/apply-for-access/. Author-generated code to support the manuscript is available on GitHub (https://doi.org/10.5281/zenodo.18979068).

## Notes

### Author Declarations

The analysis was conducted using data from UK Biobank. The UK Biobank study was approved by the North-West Multi-centre Research Ethics Committee and all participants provided written informed consent. This research has been conducted under UK Biobank project number 128619.

### Summary of Updates

Added omic only models to methods and results and supplementary. Figure 1 revised.

