## Supplementary material for "Using multiomic data to predict postoperative complications after major surgery in the UK Biobank cohort"

### Supplementary methods 1: phenotype definition and exclusions

UK Biobank (UKB) contains linked Hospital Episode Statistics (HES) inpatient data for participants. The diagnosis table contains all diagnoses (ICD-10 code) associated with each admission episode. Individuals with a previous diagnosis of the outcome phenotype of interest were excluded to reduce potential bias through confounding and reverse causation. It also ensured all events occurred after baseline variables were measured at UKB enrolment and contributed to a more homogenous case-control cohort.

The operation table contains details of all OPCS4 procedure codes for a patient during a given admission. Inpatient surgery was defined according to patient class (excluding day case). Major surgery was defined by using a combination of two grading systems from the literature (Bupa schedule of procedures and Abbott). Eligible procedures were either Bupa ‘major’ or ‘complex’ category or, if not classified in Bupa, in the ‘restrictive’ category as defined by Abbott. Only procedures performed after enrolment in UKB were eligible. All surgical specialties were included except for the acute myocardial infarction cohort, where cardiothoracic and cardiology procedures were excluded. Day case procedures were excluded based on patient classification (‘daycase’).

In cases of multiple procedures on the same date, the procedure coded as level 1 was treated as the index procedure. If there were multiple or no level 1 procedures, the procedure with the highest grade was used (complex > major > restrictive). If there were multiple procedures with the highest grade, minimum array index was used as a tiebreak. Count variables were added to account for a) multiple surgical procedures within the same hospital admission and b) cumulative procedures since UKB enrolment.

A limitation of HES coding is that procedures have a specific date of operation, whereas diagnoses are linked to an entire inpatient episode (without a specific date of diagnosis). To limit the risk of a diagnosis preceding an operative procedure where both occurred in the same inpatient episode, the following decision tree was used:

Complication diagnosis in later episode than operation date?

Yes: include

No

Operation and diagnosis in same episode. Time from episode start to operation <14 days?

Yes

Operation date in first half of episode?

Yes: include

No: exclude

No: exclude

### Supplementary methods 2: Autoencoder architectural configurations for different omic feature sets

| Input dimension (*D*) | Feature sets | Encoder architecture (neurons per layer) | Bottleneck dimension | Dropout rate | Total layers (encoder + bottleneck + decoder) |
| --- | --- | --- | --- | --- | --- |
| *D* < 512 | Metabolomics Proteomics (Infl 1) | 128, 64 | 16 | N/A | 5 |
| 512 ≤ *D* < 1024 | Proteomics (Infl 1+2)  Multiomics (Infl 1)  Multiomics (Infl 1+2) | 512, 256, 128, 64 | 16 | 0.2 | 9 |
| *D* ≥ 1024 | Proteomics (All proteins)  Multiomics (All proteins) | 1024, 512, 256, 128, 64 | 16 | 0.2 | 11 |

NB: All architectures are symmetrical: the decoder mirrors the encoder layers in reverse order. Hidden layers utilised ReLU activation; bottleneck and output layers utilised linear activation. Where present, dropout layers were added between hidden and bottleneck layers at the rate specified.

For example, the *D* < 512 architecture is:

Input layer (*D*)

|

Encoder hidden layer (128)

|

Encoder hidden layer (64)

|

Bottleneck layer (16)

|

Decoder hidden layer (64)

|

Decoder hidden layer (128)

|

Output layer (*D*)

### Supplementary methods 3: Hyperparameter grid using in Elastic Net model development

| Hyperparameter | Description | Search Space (Grid) |
| --- | --- | --- |
| C | Inverse of regularization strength (smaller values specify stronger regularization) | 0.01,0.1,1,10 |
| l1_ratio | ElasticNet mixing parameter, ranging from 0 (L2 penalty) to 1 (L1 penalty) | 0.1,0.5,0.9 |

### Supplementary methods 4: transfer learning

Conceptually, the source domain (non-postoperative) models are of the form:

*Paired phenotype ~ Age + Sex + CCI + Omics*

*For cases, age and CCI measured at the time of hospital admission related to the diagnosis. For controls, at the time of follow-up duration assigned by incidence density sampling.

The source model was then used in the postoperative cohort to generate a predicted probability (0-1) of the non-postoperative outcome:

*Predicted probability [paired phenotype] ~ Age (at time of surgery) + Sex + CCI (at time of surgery) + Omics*

The resulting predicted probabilities were then used as a feature in a parsimonious model for the postoperative outcome of interest:

*Postoperative complication ~ Predicted probability of paired phenotype + Admission Type (Elective/Emergency) + Operative Category (Major/Complex)*

This overcomes the curse of dimensionality as the number of input features for the postoperative model, in the smaller surgical cohort, has been reduced from hundreds/thousands to only three.

***Schematic of transfer learning***

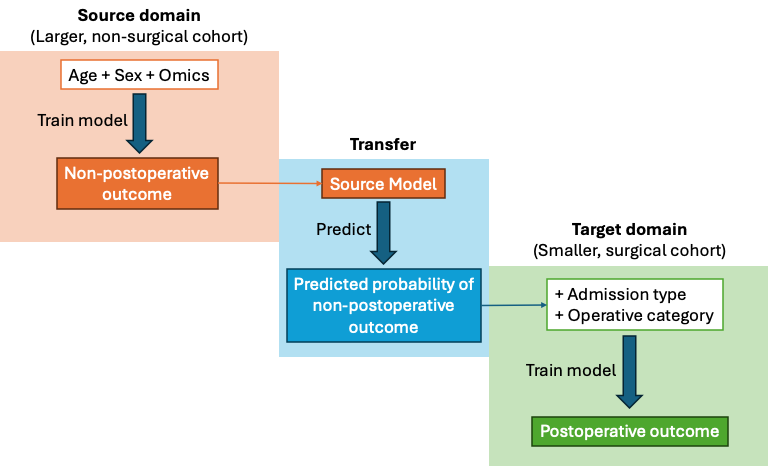

The required sample size for each resulting model is shown below (numbers are cases / total sample size):

|  | **AF** | **AKI** | **AMI** | **Delirium** | **Stroke** |
| --- | --- | --- | --- | --- | --- |
| Metabolomics | 28 / 1583 | 13 / 679 | 22 / 11725 | 26 / 3319 | 23 / 4895 |
| Proteomics | 28 / 1583 | 13 / 679 | 22 / 11725 | 26 / 3319 | 23 / 4895 |
| Multiomic | 28 / 1583 | 13 / 679 | 22 / 11725 | 26 / 3319 | 23 / 4895 |

### Supplementary methods 5: sample size analysis

**Sample size**

*Pmsampsize* (<https://cran.r-project.org/package=pmsampsize>) computes the minimum sample size required for the development of a new multivariable prediction model using the criteria proposed by Riley et al, 2018 (<https://doi.org/10.1002/sim.7992>):

i) small overfitting defined by an expected shrinkage of predictor effects by 10% or less,
ii) small absolute difference of 0.05 in the model's apparent and adjusted Nagelkerke's R-squared value, and
iii) precise estimation (within +/- 0.05) of the average outcome risk in the population for a key timepoint of interest for prediction.

The required inputs and values used are as follows:

- **C-statistic**: the C-statistic reported in an existing prediction model study, taken from the sources below:

| **Outcome** | **Citation** | **C-statistic** |
| --- | --- | --- |
| **AF** | [**https://doi.org/10.1016/j.athoracsur.2019.07.084**](https://doi.org/10.1016/j.athoracsur.2019.07.084) | **0.76** |
| **AKI** | [**https://doi.org/10.1093/ndt/gfv415**](https://doi.org/10.1093/ndt/gfv415) | **0.852 (average)** |
| **AMI** | [**https://doi.org/10.1016/j.mayocp.2019.03.008**](https://doi.org/10.1016/j.mayocp.2019.03.008) | **0.788 (average)** |
| **Delirium** | [**https://doi.org/10.1111/jgs.13138**](https://doi.org/10.1111/jgs.13138) | **0.766 (average)** |
| **SSI** | [**https://doi.org/10.1371/journal.pone.0312968**](https://doi.org/10.1371/journal.pone.0312968) | **0.79 (average, externally validated)** |
| **Stroke** | [**https://doi.org/10.3389/fnagi.2022.897611**](https://doi.org/10.3389/fnagi.2022.897611) | **0.78** |

- **Prevalence**: the outcome proportion expected within the model development dataset. Calculated based on the metabolomic sample size for each outcome.
- **Parameters**: the number of candidate predictor parameters for potential inclusion in the new model. For the different feature sets studied:
  - Metabolomics: 30 (5 clinical, 9 spectrometer, 16 autoencoder dimensions)
  - Proteomics: 21 (5 clinical, 16 autoencoder dimensions)
  - Multiomic, early integration: 30 (5 clinical, 9 spectrometer, 16 autoencoder dimensions)
  - Multiomic, late integration: 46 (5 clinical, 9 spectrometer, 16+16 autoencoder dimensions)

**Results: required number of events / total sample size for each dataset and complication**

|  | **AF** | **AKI** | **AMI** | **Delirium** | **SSI** | **Stroke** |
| --- | --- | --- | --- | --- | --- | --- |
| Metabolomics | 276 / 15826 | 130 / 6790 | 223 / 117242 | 262 / 33184 | 211 / 21110 | 229 / 48941 |
| Proteomics | 194 / 11079 | 91 / 4753 | 156 / 82069 | 183 / 23229 | 148 / 14777 | 160 / 34259 |
| Multiomic – early integration | 276 / 15826 | 130 / 6790 | 223 / 117242 | 262 / 33184 | 211 / 21110 | 229 / 48941 |
| Multiomic – late integration | 424 / 24267 | 199 / 10411 | 342 / 179770 | 401 / 50881 | 323 / 32368 | 351 / 75043 |

### Supplementary table 1: Numbers of cases and controls for each outcome and feature set

| **Dataset** | **Atrial fibrillation** | | **Acute kidney injury** | | **Acute myocardial infarction** | |
| --- | --- | --- | --- | --- | --- | --- |
|  | **Cases** | **Controls** | **Cases** | **Controls** | **Cases** | **Controls** |
| All cases | 2,610 | - | 3,067 | - | 308 | - |
| Metabolomics | 1,457 | 81,952 | 1,596 | 81,695 | 144 | 75,525 |
| Inflammation I | 242 | 14,923 | 340 | 14,741 | 31 | 13,669 |
| Inflammation I+II | 204 | 13,249 | 289 | 13,135 | 27 | 12,101 |
| All proteins | 210 | 13,418 | 296 | 13,301 | 27 | 12,261 |
| Metabolomics plus Inflammation I | 160 | 9,169 | 219 | 9,043 | 17 | 8,413 |
| Metabolomics plus inflammation I+II | 127 | 7,524 | 171 | 7,459 | 15 | 6,899 |
| Metabolomics plus all proteins | 130 | 7,543 | 172 | 7,481 | 15 | 6,924 |

| **Dataset** | **Delirium** | | **Surgical site infection** | | **Stroke** | |
| --- | --- | --- | --- | --- | --- | --- |
|  | **Cases** | **Controls** | **Cases** | **Controls** | **Cases** | **Controls** |
| All cases | 978 | - | 1,527 | - | 641 | - |
| Metabolomics | 496 | 62,367 | 838 | 83,066 | 290 | 61,733 |
| Inflammation I | 104 | 11,401 | 149 | 15,099 | 50 | 11,270 |
| Inflammation I+II | 81 | 10,106 | 129 | 13,416 | 46 | 9,982 |
| All proteins | 81 | 10,246 | 131 | 13,587 | 46 | 10,121 |
| Metabolomics plus Inflammation I | 65 | 7,067 | 94 | 9,291 | 36 | 6,976 |
| Metabolomics plus inflammation I+II | 50 | 5,764 | 76 | 7,623 | 28 | 5,692 |
| Metabolomics plus all proteins | 49 | 5,779 | 77 | 7,643 | 28 | 5,706 |

### Supplementary table 2: Post-hoc assessment of case-control numbers

Values are Pass / Fail (% required cases, % required total sample size) compared to required sample sizes detailed above (Supplementary Methods 2).

Key: >100% for both; >50% for both; 25-50% for both; <25% for both.

#### Baseline models

|  | **AF** | **AKI** | **AMI** | **Delirium** | **SSI** | **Stroke** |
| --- | --- | --- | --- | --- | --- | --- |
| **Metabolomics** | Pass | Pass | Pass | Pass | Pass | Pass |
| **Inflammation I** | Pass | Pass | Fail  (83%, 70%) | Pass | Pass | Pass |
| **Inflammation I+II** | Pass | Pass | Fail  (73%, 62%) | Pass | Pass | Pass |
| **All proteins** | Pass | Pass | Fail  (73%, 63%) | Pass | Pass | Pass |
| **Metabolomics plus Inflammation I** | Pass | Pass | Fail  (46%, 43%) | Pass | Pass | Fail  (94%, 86%) |
| **Metabolomics plus inflammation I+II** | Pass | Pass | Fail  (40%, 35%) | Pass | Pass | Fail  (73%, 70%) |
| **Metabolomics plus all proteins** | Pass | Pass | Fail  (40%, 35%) | Pass | Pass | Fail  (73%, 70%) |

#### Omic Models

|  | **AF** | **AKI** | **AMI** | **Delirium** | **SSI** | **Stroke** |
| --- | --- | --- | --- | --- | --- | --- |
| **Metabolomics** | Pass | Pass | Fail  (65%, 65%) | Pass | Pass | Pass |
| **Inflammation I** | Pass | Pass | Fail  (20%, 17%) | Fail  (57%, 50%) | Pass | Fail  (31%, 33%) |
| **Inflammation I+II** | Pass | Pass | Fail  (17%, 15%) | Fail  (44%, 44%) | Fail  (87%, 92%) | Fail  (29%, 29%) |
| **All proteins** | Pass | Pass | Fail  (17%, 15%) | Fail  (44%, 44%) | Fail  (89%, 93%) | Fail  (29%, 30%) |
| **Metabolomics plus Inflammation I, early** | Fail  (58%, 59%) | Pass | Fail  (8%, 7%) | Fail  (25%, 21%) | Fail  (45%, 44%) | Fail  (16%, 14%) |
| **Metabolomics plus inflammation I+II, early** | Fail  (46%, 48%) | Pass | Fail  (7%, 6%) | Fail  (19%, 18%) | Fail  (36%, 36%) | Fail  (12%, 12%) |
| **Metabolomics plus all proteins, early** | Fail  (47%, 48%) | Pass | Fail  (7%, 6%) | Fail  (19%, 18%) | Fail  (37%, 37%) | Fail  (12%, 12%) |
| **Metabolomics plus Inflammation I, late** | Fail  (38%, 38%) | Fail  (110%, 89%) | Fail  (5%, 5%) | Fail  (16%, 14%) | Fail  (29%, 29%) | Fail  (10%, 9%) |
| **Metabolomics plus inflammation I+II, late** | Fail  (30%, 32%) | Fail  (86%, 73%) | Fail  (4%, 4%) | Fail  (12%, 11%) | Fail  (24%, 24%) | Fail  (8%, 8%) |
| **Metabolomics plus all proteins, late** | Fail  (31%, 32%) | Fail  (86%, 74%) | Fail  (4%, 4%) | Fail  (12%, 11%) | Fail  (24%, 24%) | Fail  (8%, 8%) |

### Supplementary Table 3: Results from omic-only modelling

Values are mean AUC (standard deviation)

| **Feature set** | **Model** | **AF** | **AKI** | **AMI** | **Delirium** | **SSI** | **Stroke** |
| --- | --- | --- | --- | --- | --- | --- | --- |
| **Metabolomics** | **postoperative** | 0.57 (0.01) | 0.63 (0.01) | 0.60 (0.04) | 0.57 (0.03) | 0.56 (0.02) | 0.54 (0.04) |
|  | **non-postoperative** | 0.60 (0.01) | 0.68 (0.01) | 0.62 (0.02) | 0.61 (0.01) | 0.59 (0.01) | 0.57 (0.01) |
|  | **difference** | 0.03 | 0.05 | 0.02 | 0.05 | 0.03 | 0.04 |
| **Proteomics: infl I** | **postoperative** | 0.62 (0.04) | 0.67 (0.03) | 0.53 (0.06) | 0.63 (0.05) | 0.56 (0.06) | 0.54 (0.05) |
|  | **non-postoperative** | 0.69 (0.02) | 0.78 (0.02) | 0.66 (0.03) | 0.64 (0.03) | 0.63 (0.02) | 0.62 (0.03) |
|  | **difference** | 0.07 | 0.12 | 0.13 | 0.00 | 0.07 | 0.08 |
| **Proteomics: infl I+II** | **postoperative** | 0.55 (0.04) | 0.59 (0.03) | 0.56 (0.10) | 0.54 (0.04) | 0.54 (0.05) | 0.47 (0.06) |
|  | **non-postoperative** | 0.63 (0.02) | 0.74 (0.02) | 0.64 (0.04) | 0.59 (0.04) | 0.61 (0.02) | 0.56 (0.04) |
|  | **difference** | 0.08 | 0.15 | 0.08 | 0.05 | 0.07 | 0.09 |
| **Proteomics: all** | **postoperative** | 0.55 (0.04) | 0.58 (0.04) | 0.50 (0.08) | 0.54 (0.04) | 0.51 (0.05) | 0.45 (0.07) |
|  | **non-postoperative** | 0.60 (0.02) | 0.72 (0.02) | 0.63 (0.04) | 0.56 (0.04) | 0.56 (0.04) | 0.54 (0.04) |
|  | **difference** | 0.06 | 0.14 | 0.13 | 0.02 | 0.07 | 0.09 |
| **Multiomics: infl I** | **postoperative** | 0.59 (0.04) | 0.66 (0.03) | 0.55 (0.10) | 0.52 (0.05) | 0.52 (0.04) | 0.46 (0.07) |
|  | **non-postoperative** | 0.65 (0.02) | 0.77 (0.02) | 0.60 (0.05) | 0.61 (0.04) | 0.61 (0.03) | 0.61 (0.03) |
|  | **difference** | 0.06 | 0.11 | 0.05 | 0.09 | 0.09 | 0.15 |
| **Multiomics: infl I+II** | **postoperative** | 0.54 (0.04) | 0.61 (0.04) | 0.59 (0.09) | 0.58 (0.07) | 0.51 (0.05) | 0.46 (0.08) |
|  | **non-postoperative** | 0.64 (0.03) | 0.74 (0.02) | 0.58 (0.06) | 0.59 (0.04) | 0.59 (0.03) | 0.58 (0.04) |
|  | **difference** | 0.09 | 0.13 | -0.01 | 0.01 | 0.08 | 0.12 |
| **Multiomics: all proteins** | **postoperative** | 0.57 (0.04) | 0.58 (0.04) | 0.59 (0.10) | 0.50 (0.06) | 0.49 (0.04) | 0.45 (0.08) |
|  | **non-postoperative** | 0.60 (0.03) | 0.72 (0.03) | 0.59 (0.05) | 0.56 (0.06) | 0.59 (0.03) | 0.57 (0.04) |
|  | **difference** | 0.03 | 0.14 | 0.00 | 0.05 | 0.09 | 0.12 |

### Supplementary table 4: Predictive model evaluation results

P-values calculated using Nadeau & Bengio’s Corrected Resampled t-test to account for data overlap in repeated 3/5-fold CV. The test incorporates a correction factor based on the ratio of test-to-training data (1/*k*-1) to adjust the t-statistic accordingly. Multiple testing handled via Benjamini-Hochberg FDR correction applied per comparison type.

**Supplementary Table 4.1** Model performance across different feature sets for **postoperative acute kidney injury**. Values are mean ± standard deviation. For multiomic feature sets, all include metabolomics and metrics shown are for early integration models. AUROC: area under the receiver operator characteristic curve.

|  | AUROC |  |  | Sensitivity |  |  | Specificity |  |  |
| --- | --- | --- | --- | --- | --- | --- | --- | --- | --- |
| Feature Set | Baseline | + Omics | Transfer | Baseline | + Omics | Transfer | Baseline | + Omics | Transfer |
| Metabolomics | 0.81 ± 0.01 | 0.82 ± 0.01** | 0.82 ± 0.01 | 0.72 ± 0.03 | 0.72 ± 0.02 | 0.72 ± 0.03 | 0.75 ± 0.00 | 0.76 ± 0.00*** | 0.76 ± 0.00^ |
| Inflammation I | 0.81 ± 0.02 | 0.82 ± 0.02 | 0.82 ± 0.02 | 0.72 ± 0.05 | 0.72 ± 0.05 | 0.72 ± 0.04 | 0.75 ± 0.01 | 0.77 ± 0.01*** | 0.76 ± 0.01 |
| Inflammation I+II | 0.81 ± 0.03 | 0.81 ± 0.02 | 0.81 ± 0.02 | 0.71 ± 0.06 | 0.72 ± 0.05 | 0.72 ± 0.05 | 0.75 ± 0.01 | 0.76 ± 0.01 | 0.75 ± 0.01 |
| All Proteins | 0.81 ± 0.02 | 0.81 ± 0.02 | 0.81 ± 0.02 | 0.71 ± 0.06 | 0.73 ± 0.05 | 0.73 ± 0.05 | 0.75 ± 0.01 | 0.76 ± 0.01 | 0.74 ± 0.01 |
| Multiomic  (Inflammation I) | 0.81 ± 0.03 | 0.82 ± 0.03 | 0.81 ± 0.03 | 0.71 ± 0.06 | 0.72 ± 0.05 | 0.70 ± 0.06 | 0.76 ± 0.01 | 0.77 ± 0.01 | 0.76 ± 0.01 |
| Multiomic  (Inflammation I+II) | 0.81 ± 0.03 | 0.80 ± 0.03 | 0.81 ± 0.03 | 0.71 ± 0.07 | 0.71 ± 0.07 | 0.71 ± 0.07 | 0.76 ± 0.01 | 0.76 ± 0.01 | 0.76 ± 0.01 |
| Multiomic  (All Proteins) | 0.81 ± 0.03 | 0.80 ± 0.04 | 0.81 ± 0.03 | 0.71 ± 0.08 | 0.71 ± 0.08 | 0.69 ± 0.06 | 0.76 ± 0.01 | 0.76 ± 0.01 | 0.76 ± 0.01 |

*adjusted p-value Model vs Baseline < 0.05; **adjusted p-value Model vs Baseline < 0.01; ***adjusted p-value Model vs Baseline < 0.001

^ adjusted p-value Transfer vs Baseline < 0.05; ^^adjusted p-value Transfer vs Baseline < 0.01; ^^^ adjusted p-value Transfer vs Baseline < 0.001

$ adjusted p-value Transfer vs Model < 0.05; $$ adjusted p-value Transfer vs Model < 0.01; $$$ adjusted p-value Transfer vs Model < 0.001

**Supplementary Table 4.2** Model performance across different feature sets for **postoperative atrial fibrillation**. Values are mean ± standard deviation. For multiomic feature sets, all include metabolomics and metrics shown are for early integration models. AUROC: area under the receiver operator characteristic curve.

|  | AUROC |  |  | Sensitivity |  |  | Specificity |  |  |
| --- | --- | --- | --- | --- | --- | --- | --- | --- | --- |
| Feature Set | Baseline | + Omics | Transfer | Baseline | + Omics | Transfer | Baseline | + Omics | Transfer |
| Metabolomics | 0.83 ± 0.01 | 0.83 ± 0.01 | 0.82 ± 0.01 | 0.79 ± 0.02 | 0.78 ± 0.02 | 0.77 ± 0.03 | 0.73 ± 0.00 | 0.73 ± 0.00** | 0.73 ± 0.00 |
| Inflammation I | 0.84 ± 0.02 | 0.83 ± 0.03 | 0.83 ± 0.02 | 0.78 ± 0.07 | 0.77 ± 0.06 | 0.78 ± 0.06 | 0.74 ± 0.01 | 0.75 ± 0.01** | 0.73 ± 0.01$ |
| Inflammation I+II | 0.83 ± 0.03 | 0.83 ± 0.02 | 0.83 ± 0.02 | 0.78 ± 0.07 | 0.76 ± 0.06 | 0.78 ± 0.06 | 0.74 ± 0.01 | 0.74 ± 0.01 | 0.73 ± 0.01 |
| All Proteins | 0.83 ± 0.03 | 0.82 ± 0.03 | 0.84 ± 0.02 | 0.77 ± 0.07 | 0.76 ± 0.06 | 0.79 ± 0.07 | 0.74 ± 0.01 | 0.73 ± 0.01 | 0.74 ± 0.01 |
| Multiomic  (Inflammation I) | 0.85 ± 0.02 | 0.84 ± 0.03 | 0.81 ± 0.02 | 0.77 ± 0.06 | 0.75 ± 0.06 | 0.73 ± 0.07 | 0.75 ± 0.01 | 0.75 ± 0.01 | 0.71 ± 0.01^^^$$$ |
| Multiomic  (Inflammation I+II) | 0.84 ± 0.03 | 0.83 ± 0.03 | 0.81 ± 0.03 | 0.76 ± 0.08 | 0.73 ± 0.08 | 0.75 ± 0.07 | 0.75 ± 0.02 | 0.75 ± 0.01 | 0.70 ± 0.01^^^$$$ |
| Multiomic  (All Proteins) | 0.84 ± 0.03 | 0.83 ± 0.03 | 0.81 ± 0.03 | 0.76 ± 0.09 | 0.74 ± 0.09 | 0.69 ± 0.08 | 0.75 ± 0.02 | 0.75 ± 0.02 | 0.73 ± 0.01 |

*adjusted p-value Model vs Baseline < 0.05; **adjusted p-value Model vs Baseline < 0.01; ***adjusted p-value Model vs Baseline < 0.001

^ adjusted p-value Transfer vs Baseline < 0.05; ^^adjusted p-value Transfer vs Baseline < 0.01; ^^^ adjusted p-value Transfer vs Baseline < 0.001

$ adjusted p-value Transfer vs Model < 0.05; $$ adjusted p-value Transfer vs Model < 0.01; $$$ adjusted p-value Transfer vs Model < 0.001

**Supplementary Table 4.3** Model performance across different feature sets for **postoperative delirium**. Values are mean ± standard deviation. For multiomic feature sets, all include metabolomics and metrics shown are for early integration models. AUROC: area under the receiver operator characteristic curve.

|  | AUROC |  |  | Sensitivity |  |  | Specificity |  |  |
| --- | --- | --- | --- | --- | --- | --- | --- | --- | --- |
| Feature Set | Baseline | + Omics | Transfer | Baseline | + Omics | Transfer | Baseline | + Omics | Transfer |
| Metabolomics | 0.88 ± 0.02 | 0.87 ± 0.02 | 0.87 ± 0.02 | 0.79 ± 0.04 | 0.78 ± 0.05 | 0.78 ± 0.04 | 0.80 ± 0.00 | 0.80 ± 0.00* | 0.80 ± 0.01 |
| Inflammation I | 0.85 ± 0.04 | 0.84 ± 0.03 | 0.86 ± 0.04 | 0.79 ± 0.08 | 0.78 ± 0.08 | 0.80 ± 0.07 | 0.80 ± 0.01 | 0.80 ± 0.01 | 0.79 ± 0.01 |
| Inflammation I+II | 0.83 ± 0.04 | 0.80 ± 0.05 | 0.86 ± 0.04 | 0.76 ± 0.08 | 0.70 ± 0.09 | 0.79 ± 0.07 | 0.79 ± 0.02 | 0.79 ± 0.02 | 0.79 ± 0.01 |
| All Proteins | 0.83 ± 0.03 | 0.80 ± 0.04 | 0.85 ± 0.04 | 0.76 ± 0.06 | 0.71 ± 0.07 | 0.76 ± 0.08 | 0.79 ± 0.01 | 0.77 ± 0.02 | 0.79 ± 0.01 |
| Multiomic  (Inflammation I) | 0.86 ± 0.04 | 0.82 ± 0.04 | 0.82 ± 0.03 | 0.82 ± 0.07 | 0.75 ± 0.08 | 0.76 ± 0.08 | 0.81 ± 0.01 | 0.80 ± 0.02 | 0.76 ± 0.01^^ |
| Multiomic  (Inflammation I+II) | 0.84 ± 0.05 | 0.78 ± 0.06 | 0.79 ± 0.03 | 0.81 ± 0.08 | 0.69 ± 0.11 | 0.71 ± 0.07 | 0.80 ± 0.01 | 0.79 ± 0.02 | 0.77 ± 0.01^ |
| Multiomic  (All Proteins) | 0.84 ± 0.06 | 0.78 ± 0.07 | 0.78 ± 0.04 | 0.81 ± 0.10 | 0.70 ± 0.12 | 0.68 ± 0.08 | 0.80 ± 0.02 | 0.78 ± 0.03 | 0.76 ± 0.02^ |

*adjusted p-value Model vs Baseline < 0.05; **adjusted p-value Model vs Baseline < 0.01; ***adjusted p-value Model vs Baseline < 0.001

^ adjusted p-value Transfer vs Baseline < 0.05; ^^adjusted p-value Transfer vs Baseline < 0.01; ^^^ adjusted p-value Transfer vs Baseline < 0.001

$ adjusted p-value Transfer vs Model < 0.05; $$ adjusted p-value Transfer vs Model < 0.01; $$$ adjusted p-value Transfer vs Model < 0.001

**Supplementary Table 4.4** Model performance across different feature sets for **postoperative myocardial infarction**. Values are mean ± standard deviation. For multiomic feature sets, all include metabolomics and metrics shown are for early integration models. AUROC: area under the receiver operator characteristic curve.

|  | AUROC |  |  | Sensitivity |  |  | Specificity |  |  |
| --- | --- | --- | --- | --- | --- | --- | --- | --- | --- |
| Feature Set | Baseline | + Omics | Transfer | Baseline | + Omics | Transfer | Baseline | + Omics | Transfer |
| Metabolomics | 0.85 ± 0.03 | 0.84 ± 0.04 | 0.86 ± 0.03 | 0.76 ± 0.08 | 0.73 ± 0.09 | 0.78 ± 0.06 | 0.79 ± 0.01 | 0.80 ± 0.01** | 0.79 ± 0.01 |
| Inflammation I | 0.87 ± 0.04 | 0.79 ± 0.06 | 0.87 ± 0.05 | 0.81 ± 0.09 | 0.62 ± 0.17 | 0.82 ± 0.12 | 0.80 ± 0.02 | 0.80 ± 0.02 | 0.80 ± 0.02 |
| Inflammation I+II | 0.88 ± 0.06 | 0.80 ± 0.08 | 0.89 ± 0.04 | 0.80 ± 0.15 | 0.65 ± 0.17 | 0.79 ± 0.11 | 0.80 ± 0.02 | 0.80 ± 0.02 | 0.81 ± 0.02 |
| All Proteins | 0.89 ± 0.03 | 0.72 ± 0.09* | 0.87 ± 0.05 | 0.82 ± 0.11 | 0.63 ± 0.15 | 0.82 ± 0.11 | 0.81 ± 0.01 | 0.77 ± 0.02 | 0.81 ± 0.02 |
| Multiomic (Infl I) | 0.83 ± 0.08 | 0.60 ± 0.11* | 0.85 ± 0.07 | 0.82 ± 0.13 | 0.44 ± 0.21 | 0.75 ± 0.13 | 0.80 ± 0.01 | 0.70 ± 0.04** | 0.80 ± 0.01$ |
| Multiomic (Infl I+II) | 0.82 ± 0.10 | 0.65 ± 0.10 | 0.86 ± 0.06 | 0.78 ± 0.17 | 0.46 ± 0.20 | 0.76 ± 0.17 | 0.80 ± 0.02 | 0.75 ± 0.03 | 0.80 ± 0.01 |
| Multiomic (All Prot) | 0.81 ± 0.10 | 0.66 ± 0.09 | 0.84 ± 0.07 | 0.76 ± 0.20 | 0.53 ± 0.21 | 0.83 ± 0.12 | 0.80 ± 0.02 | 0.70 ± 0.05* | 0.80 ± 0.01$ |

*adjusted p-value Model vs Baseline < 0.05; **adjusted p-value Model vs Baseline < 0.01; ***adjusted p-value Model vs Baseline < 0.001

^ adjusted p-value Transfer vs Baseline < 0.05; ^^adjusted p-value Transfer vs Baseline < 0.01; ^^^ adjusted p-value Transfer vs Baseline < 0.001

$ adjusted p-value Transfer vs Model < 0.05; $$ adjusted p-value Transfer vs Model < 0.01; $$$ adjusted p-value Transfer vs Model < 0.001

**Supplementary Table 4.5** Model performance across different feature sets for **surgical site infection**. Values are mean ± standard deviation. For multiomic feature sets, all include metabolomics and metrics shown are for early integration models. AUROC: area under the receiver operator characteristic curve.

|  | AUROC |  |  | Sensitivity |  |  | Specificity |  |  |
| --- | --- | --- | --- | --- | --- | --- | --- | --- | --- |
| Feature Set | Baseline | + Omics | Transfer | Baseline | + Omics | Transfer | Baseline | + Omics | Transfer |
| Metabolomics | 0.72 ± 0.02 | 0.73 ± 0.02 | 0.71 ± 0.02 | 0.75 ± 0.03 | 0.72 ± 0.03* | 0.64 ± 0.04***$ | 0.65 ± 0.00 | 0.67 ± 0.01*** | 0.68 ± 0.01^^^ |
| Inflammation I | 0.71 ± 0.04 | 0.72 ± 0.04 | 0.72 ± 0.04 | 0.74 ± 0.08 | 0.64 ± 0.09* | 0.62 ± 0.08 | 0.64 ± 0.02 | 0.70 ± 0.01*** | 0.70 ± 0.01^^^ |
| Inflammation I+II | 0.72 ± 0.04 | 0.73 ± 0.04 | 0.72 ± 0.04 | 0.79 ± 0.08 | 0.68 ± 0.09* | 0.66 ± 0.09 | 0.64 ± 0.01 | 0.69 ± 0.01*** | 0.69 ± 0.01^^^ |
| All Proteins | 0.72 ± 0.04 | 0.71 ± 0.06 | 0.72 ± 0.04 | 0.78 ± 0.07 | 0.69 ± 0.09* | 0.64 ± 0.08* | 0.64 ± 0.01 | 0.68 ± 0.01*** | 0.69 ± 0.01^^^ |
| Multiomic (Infl I) | 0.73 ± 0.04 | 0.69 ± 0.04 | 0.68 ± 0.04 | 0.73 ± 0.07 | 0.61 ± 0.08 | 0.53 ± 0.07* | 0.67 ± 0.02 | 0.71 ± 0.01* | 0.74 ± 0.02^^ |
| Multiomic (Infl I+II) | 0.73 ± 0.03 | 0.69 ± 0.03 | 0.68 ± 0.04 | 0.76 ± 0.07 | 0.61 ± 0.09* | 0.55 ± 0.06* | 0.66 ± 0.02 | 0.71 ± 0.02*** | 0.74 ± 0.03^^ |
| Multiomic (All Prot) | 0.73 ± 0.04 | 0.69 ± 0.05 | 0.64 ± 0.04 | 0.76 ± 0.07 | 0.65 ± 0.11 | 0.52 ± 0.07* | 0.66 ± 0.02 | 0.70 ± 0.02* | 0.78 ± 0.01^^^ |

*adjusted p-value Model vs Baseline < 0.05; **adjusted p-value Model vs Baseline < 0.01; ***adjusted p-value Model vs Baseline < 0.001

^ adjusted p-value Transfer vs Baseline < 0.05; ^^adjusted p-value Transfer vs Baseline < 0.01; ^^^ adjusted p-value Transfer vs Baseline < 0.001

$ adjusted p-value Transfer vs Model < 0.05; $$ adjusted p-value Transfer vs Model < 0.01; $$$ adjusted p-value Transfer vs Model < 0.001

**Supplementary Table 4.6** Model performance across different feature sets for **postoperative stroke**. Values are mean ± standard deviation. For multiomic feature sets, all include metabolomics and metrics shown are for early integration models. AUROC: area under the receiver operator characteristic curve.

|  | AUROC |  |  | Sensitivity |  |  | Specificity |  |  |
| --- | --- | --- | --- | --- | --- | --- | --- | --- | --- |
| Feature Set | Baseline | + Omics | Transfer | Baseline | + Omics | Transfer | Baseline | + Omics | Transfer |
| Metabolomics | 0.79 ± 0.03 | 0.79 ± 0.03 | 0.79 ± 0.03 | 0.72 ± 0.05 | 0.71 ± 0.06 | 0.72 ± 0.07 | 0.77 ± 0.01 | 0.76 ± 0.01*** | 0.77 ± 0.01 |
| Inflammation I | 0.76 ± 0.04 | 0.70 ± 0.05 | 0.79 ± 0.03 | 0.69 ± 0.11 | 0.54 ± 0.12 | 0.69 ± 0.10 | 0.70 ± 0.04 | 0.74 ± 0.02 | 0.74 ± 0.01 |
| Inflammation I+II | 0.78 ± 0.04 | 0.72 ± 0.05 | 0.78 ± 0.04 | 0.70 ± 0.12 | 0.60 ± 0.11 | 0.69 ± 0.10 | 0.73 ± 0.02 | 0.74 ± 0.02 | 0.73 ± 0.01 |
| All Proteins | 0.78 ± 0.04 | 0.67 ± 0.08 | 0.77 ± 0.04 | 0.71 ± 0.11 | 0.56 ± 0.13 | 0.72 ± 0.10 | 0.72 ± 0.03 | 0.71 ± 0.02 | 0.72 ± 0.02 |
| Multiomic (Infl I) | 0.74 ± 0.05 | 0.59 ± 0.08* | 0.70 ± 0.07 | 0.63 ± 0.14 | 0.46 ± 0.12 | 0.61 ± 0.15 | 0.69 ± 0.04 | 0.71 ± 0.02 | 0.73 ± 0.03 |
| Multiomic (Infl I+II) | 0.73 ± 0.08 | 0.60 ± 0.09 | 0.69 ± 0.07 | 0.47 ± 0.31 | 0.51 ± 0.30 | 0.67 ± 0.07 | 0.73 ± 0.17 | 0.73 ± 0.11 | 0.71 ± 0.02 |
| Multiomic (All Prot) | 0.75 ± 0.07 | 0.60 ± 0.09* | 0.68 ± 0.06 | 0.57 ± 0.34 | 0.35 ± 0.28 | 0.67 ± 0.12 | 0.65 ± 0.27 | 0.80 ± 0.15 | 0.72 ± 0.02 |

*adjusted p-value Model vs Baseline < 0.05; **adjusted p-value Model vs Baseline < 0.01; ***adjusted p-value Model vs Baseline < 0.001

^ adjusted p-value Transfer vs Baseline < 0.05; ^^adjusted p-value Transfer vs Baseline < 0.01; ^^^ adjusted p-value Transfer vs Baseline < 0.001

$ adjusted p-value Transfer vs Model < 0.05; $$ adjusted p-value Transfer vs Model < 0.01; $$$ adjusted p-value Transfer vs Model < 0.001

### Supplementary table 5: Number of cases in target (postoperative) and source (non-postoperative) domains for transfer learning and ratio between the two (source divided by target)

| Complication | Dataset | Non-postoperative cases | Non-postoperative controls | Postoperative cases | Ratio |
| --- | --- | --- | --- | --- | --- |
| Atrial fibrillation | Metabolomics | 8518 | 34070 | 1457 | 5.8 |
|  | Inflammation I | 1576 | 6311 | 242 | 6.5 |
|  | Inflammation I+II | 1386 | 5628 | 204 | 6.8 |
|  | All proteins | 1392 | 5695 | 210 | 6.6 |
|  | Metabolomics plus Inflammation I | 1002 | 4017 | 160 | 6.3 |
|  | Metabolomics plus inflammation I+II | 816 | 3319 | 127 | 6.4 |
|  | Metabolomics plus all proteins | 814 | 3319 | 130 | 6.3 |
| Acute kidney injury | Metabolomics | 5431 | 21723 | 1596 | 3.4 |
|  | Inflammation I | 1156 | 4643 | 340 | 3.4 |
|  | Inflammation I+II | 1005 | 4119 | 289 | 3.5 |
|  | All proteins | 1012 | 4180 | 296 | 3.4 |
|  | Metabolomics plus Inflammation I | 726 | 2911 | 219 | 3.3 |
|  | Metabolomics plus inflammation I+II | 584 | 2430 | 171 | 3.4 |
|  | Metabolomics plus all proteins | 584 | 2442 | 172 | 3.4 |
| Acute myocardial infarction | Metabolomics | 1705 | 6824 | 144 | 11.8 |
|  | Inflammation I | 315 | 1270 | 31 | 10.2 |
|  | Inflammation I+II | 282 | 1130 | 27 | 10.4 |
|  | All proteins | 287 | 1137 | 27 | 10.6 |
|  | Metabolomics plus Inflammation I | 213 | 841 | 17 | 12.5 |
|  | Metabolomics plus inflammation I+II | 171 | 690 | 15 | 11.4 |
|  | Metabolomics plus all proteins | 172 | 689 | 15 | 11.5 |
| Delirium | Metabolomics | 1939 | 5211 | 496 | 3.9 |
|  | Inflammation I | 436 | 1195 | 104 | 4.2 |
|  | Inflammation I+II | 367 | 1053 | 81 | 4.5 |
|  | All proteins | 370 | 1059 | 81 | 4.6 |
|  | Metabolomics plus Inflammation I | 257 | 711 | 65 | 4 |
|  | Metabolomics plus inflammation I+II | 201 | 587 | 50 | 4 |
|  | Metabolomics plus all proteins | 200 | 586 | 49 | 4.1 |
| Surgical site infection | Metabolomics | 4414 | 17659 | 838 | 5.3 |
|  | Inflammation I | 854 | 3479 | 149 | 5.7 |
|  | Inflammation I+II | 750 | 3090 | 129 | 5.8 |
|  | All proteins | 758 | 3111 | 131 | 5.8 |
|  | Metabolomics plus Inflammation I | 529 | 2102 | 94 | 5.6 |
|  | Metabolomics plus inflammation I+II | 435 | 1721 | 76 | 5.7 |
|  | Metabolomics plus all proteins | 432 | 1729 | 77 | 5.6 |
| Stroke | Metabolomics | 2851 | 8213 | 290 | 9.8 |
|  | Inflammation I | 547 | 1543 | 50 | 10.9 |
|  | Inflammation I+II | 465 | 1371 | 46 | 10.1 |
|  | All proteins | 476 | 1375 | 46 | 10.3 |
|  | Metabolomics plus Inflammation I | 332 | 976 | 36 | 9.2 |
|  | Metabolomics plus inflammation I+II | 251 | 811 | 28 | 9 |
|  | Metabolomics plus all proteins | 258 | 815 | 28 | 9.2 |

### Supplementary figure 1: Feature sets and model inputs including early and late integration methods.

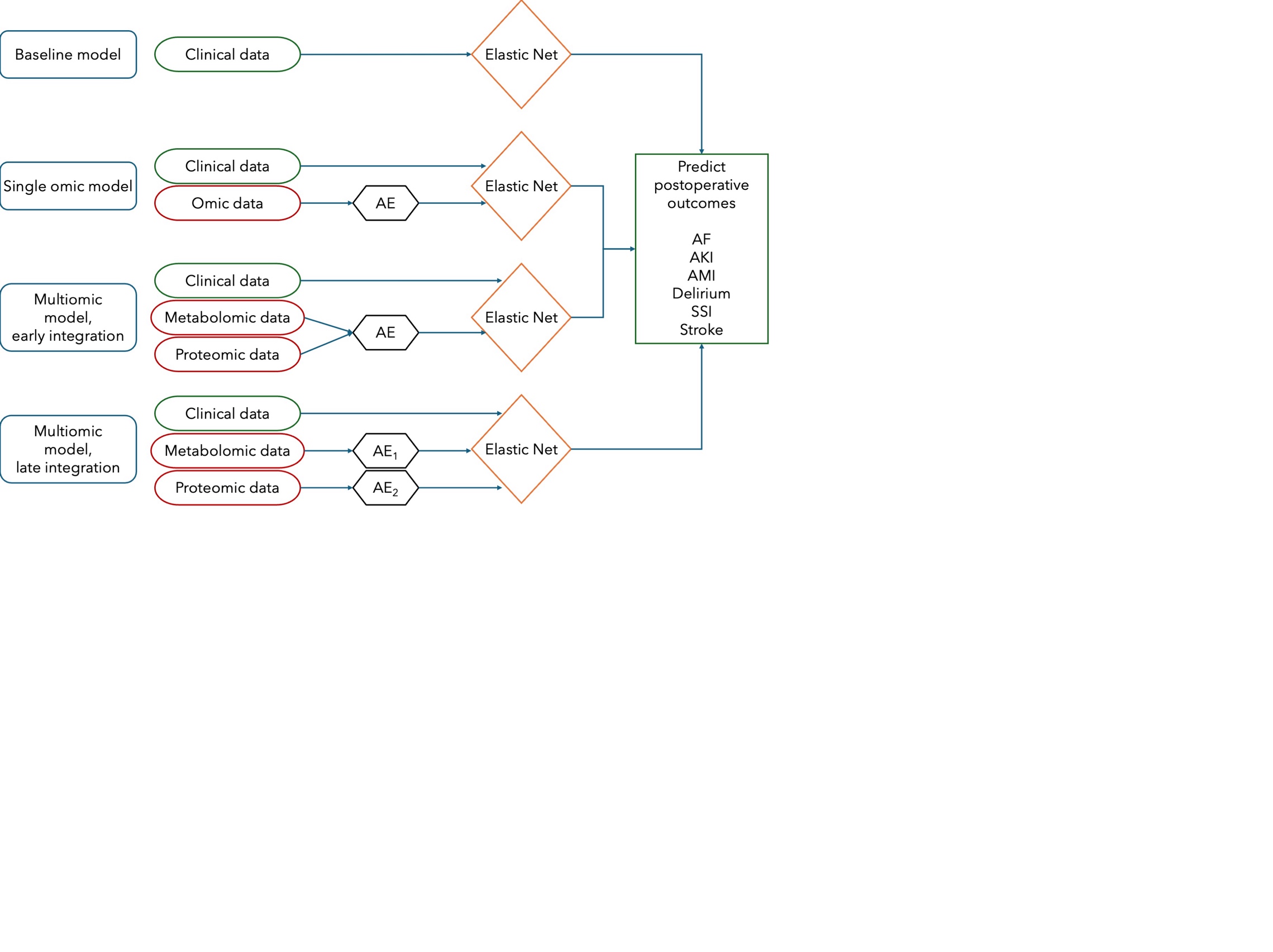

AE: autoencoder.

### Supplementary figure 2: Dynamic adaptive crossvalidation framework

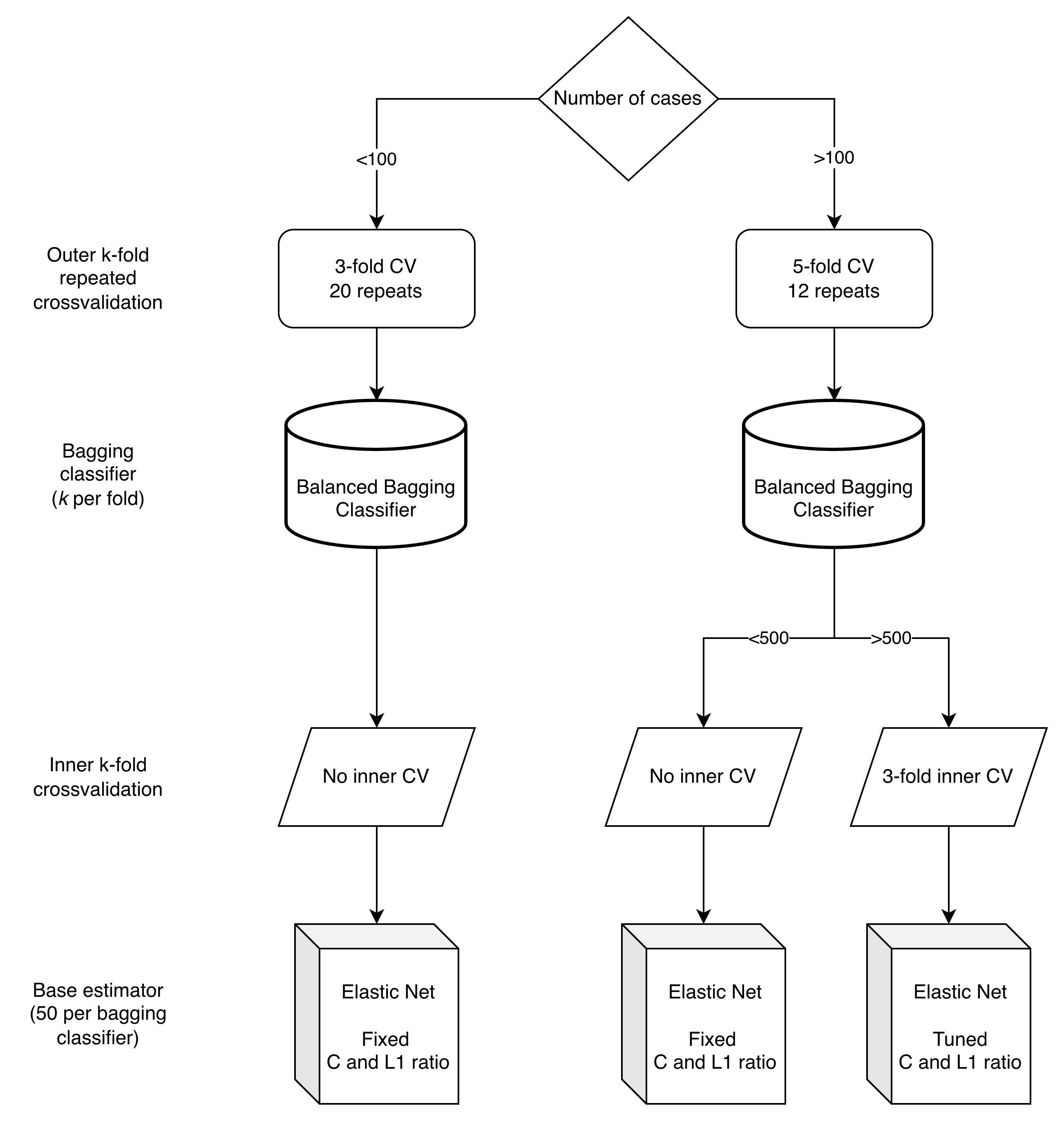
